# Differences in AD-related pathology profiles across APOE groups

**DOI:** 10.1101/2023.04.25.23289108

**Authors:** Cassandra Morrison, Mahsa Dadar, Farooq Kamal, D. Louis Collins, the Alzheimer’s Disease Neuroimaging Initiative

**Affiliations:** McConnell Brain Imaging Centre, Montreal Neurological Institute, McGill University, Montreal, Quebec, H3A 2B4, Canada; Department of Neurology and Neurosurgery, McGill University, Montreal, Quebec, H3A 2B4, Canada; Department of Psychiatry, McGill University, Montreal, Quebec, H3A 1A1, Canada; Douglas Mental Health University Institute, Montreal, Quebec, H4H 1R3, Canada

**Author notes:** **Corresponding author:** Cassandra Morrison, Montreal Neurological Institute, 3801 University Street, Montreal QC, H3A 2B4. Data used in preparation of this article were obtained from the Alzheimer’s Disease Neuroimaging Initiative (ADNI) database (adni.loni.usc.edu). As such, the investigators within the ADNI contributed to the design and implementation of ADNI and/or provided data but did not participate in analysis or writing of this report. A complete listing of ADNI investigators can be found at: http://adni.loni.usc.edu/wp-content/uploads/how_to_apply/ADNI_Acknowledgement_List.pdf.

**Keywords:** Older Adults, Pathology, White Matter Hyperintensities, APOE, Atrophy, Amyloid, Tau, Apolipoprotein

## Abstract

**BACKGROUND:** The apolipoprotein (APOE) e4 allele is a known risk factor for Alzheimer’s disease (AD), while the e2 allele is thought to be protective against AD. Few studies have examined the relationship between brain pathologies, atrophy, and white matter hyperintensities (WMHs) and APOE status in those with the e2e4 genotype and results are inconsistent for those with an e2 allele.

**METHODS:** We analyzed Alzheimer’s Disease Neuroimaging participants that had APOE genotyping and at least one of the following metrics: regional WMH load, ventricle size, hippocampal (HC) and entorhinal cortex (EC) volume, amyloid level (i.e., AV-45), and phosphorylated tau (pTau). Participants were divided into one of four APOE allele profiles (E4=e4e4 or e3e4; E2=e2e2 or e2e3; E3=e3e3; or E24=e2e4, Fig.1). Linear mixed models examined the relationship between APOE profiles and each pathology (i.e., regional WMHs, ventricle size, hippocampal and entorhinal cortex volume, amyloid level, and phosphorylated tau measures). while controlling for age, sex, education, and diagnostic status at baseline and over time.

**RESULTS:** APOE ε4 is associated with increased pathology while ε2 positivity is associated with reduced baseline and lower accumulation of pathologies and rates of neurodegeneration. APOE ε2ε4 is similar to ε4 (increased neurodegeneration) but with a slower rate of change.

**CONCLUSIONS:** The strong associations observed between APOE and pathology in this study show the importance of how genetic factors influence structural brain changes. These findings suggest that ε2ε4 genotype is related to increased declines associated with the ε4 as opposed to the protective effects of the ε2. These findings have important implications for initiating treatments and interventions. Given that people who have the ε2ε4 genotype can expect to have increased atrophy, they must be included (alongside those with an ε4 profile) in targeted interventions to reduce brain changes that occur with AD.

## 1. Background

The ε4 allele of the apolipoprotein E (APOE) gene is associated with a significant risk for the development of Alzheimer’s disease (AD) (1). While one ε4 allele has been shown to increase risk for AD by approximately 30%, two ε4 alleles increases risk by approximately 65%. Furthermore, the presence of the ε4 allele decreases the mean age of onset for AD diagnosis in a dose dependent manner (2,3), and is associated with faster disease progression compared to non-ε4 carriers (2). On the other hand, ε2 carriers are observed to exhibit up to a 50% less risk of AD compared to ε3/ε3 genotypes, and later mean age of onset (4)(4,5).

Given the relationship between APOE status and AD risk, several studies have examined the association between APOE genotype and AD pathology. Research has reported an association between ε4 and greater beta amyloid (Aβ) deposition (6). However, the relationship between ε4 and tau pathology may be more complex. While researchers have observed that the ε4 is associated with increased tau accumulation (7–9), some have reported that this relationship is observed only when Aβ is also present (10). The ε2 allele has been found to be associated with low levels of tau (11–13) and Aβ deposition (11,12). Of note, are two studies which observed the ε2ε4 genotype had similar baseline Thal phase amyloid (11), Braak staging (11,12), and Neuritic plaques (11,12) compared to that of ε4, but with less severity. Nevertheless, in most studies, the ε2 and ε4 are compared to the neutral risk ε3 alleles.

Several reviews have reported that the ε4 is associated with extensive atrophy, especially in AD-specific brain regions such as the hippocampus, amygdala, entorhinal cortex, as well as with ventricular enlargement (14,15). While some studies have reported lower atrophy rates in those with the ε2 allele compared to ε3 homozygotes (two ε3 alleles) (16,17) and those with an ε4 allele (17) these atrophy differences associated with the ε2 versus other APOE genotypes are not always observed (18). Taken together, these findings indicate that more research is needed to fully understand the relationship between APOE status and AD-specific measures of neurodegeneration.

Another contributor to AD risk is cerebral small vessel disease (CSVD) (19), which is often quantified using white matter hyperintensities (WMHs) on T2 or FLAIR MRI (20). Increased WMH burden increases cognitive decline in normal aging (21) and progression to mild cognitive impairment and dementia (20,22). Previous research has observed a significant association between the ε4 (23–25) and ε2 (23,26) and WMH burden. The relationship observed between the ε4 allele and WMH burden may explain why approximately 70% of diagnosed AD cases are of a mixed etiology (27).

To date, most research examining the influence of APOE on brain pathology, compares ε4 homozygotes (two ε4 alleles) to ε4 heterozygotes (one ε4 and one ε3 allele) and ε3 homozygotes. This type of method is important to understand the dose dependent effect of the ε4 on brain pathology, but does not provide a clear understanding of how different APOE genotypes influence brain changes. The results remain unclear whether there are differences in brain pathology in those with an ε2 compared to other APOE profiles. Most studies exclude people who exhibit the ε2ε4 genotype because of the combined protective and detrimental nature of the two alleles and because this genotype is less common than other types. It remains unknown whether people with both an ε4 and ε2 allele have increased or decreased pathology change over time relative to other APOE profiles. It is thus possible that in response to some pathologies the ε2 is protective and that the ε4 is detrimental for other pathologies. Furthermore, these studies have yet to examine whether rate of change in various pathologies differ based on APOE profile. The goal of this paper was to examine AD-related pathologies in a longitudinal manner to improve our current understanding of how these pathological mechanisms are influenced by APOE.

## 2. Methods

### 2.1 Alzheimer’s Disease Neuroimaging Initiative

Data used in the preparation of this article were obtained from the Alzheimer’s Disease Neuroimaging Initiative (ADNI) database (adni.loni.usc.edu). The ADNI was launched in 2003 as a public-private partnership, led by Principal Investigator Michael W. Weiner, MD. The primary goal of ADNI has been to test whether serial MRI, positron emission tomography (PET), other biological markers, and clinical and neuropsychological assessment can be combined to measure the progression of mild cognitive impairment and early AD. The study received ethical approval from the review boards of all participating institutions. Written informed consent was obtained from participants or their study partner. Participants were selected from all ADNI Cohorts (ADNI-1, ADNI-GO, ADNI-2 and ADNI-3).

### 2.2 Participants

Full participant inclusion and exclusion criteria are available at www.adni-info.org. All participants were between the ages of 55 and 90 at baseline, with no evidence of depression. Cognitively healthy older adults exhibited no evidence of memory decline, as measured by the Wechsler Memory Scale and no evidence of impaired global cognition as measured by the Mini Mental Status Examination (MMSE) or Clinical Dementia Rating (CDR). MCI participants scored between 24 and 30 on the MMSE, 0.5 on the CDR, and abnormal scores on the Wechsler Memory Scale. Dementia was defined as participants who had abnormal memory function on the Wechsler Memory Scale, an MMSE score between 20 and 26 and a CDR of 0.5 or 1.0 and a probable AD clinical diagnosis according to the National Institute of Neurological and Communicative Disorders and Stroke and the Alzheimer’s Disease and Related Disorders Association criteria.

Participants were included if they had completed APOE genotyping and had at least one of the dependent variables of interest. That is, information from at least one of the following: MRIs from which WMHs could be extracted, or ventricle, hippocampal, and entorhinal cortex volumes, or pTau measures, or AV-45 measures. A total of 2119 participants with 9847 timepoints with MRIs from which WMHs could be extracted were included in the WMH analysis. A total of 2050 participants with 8707 timepoints had ventricle volumes, 2006 participants with 8026 timepoints had HC volumes, and 1968 participants with 7630 had EC volumes. Only 1231 participants with 2412 timepoints had pTau measurements and 1212 participants with 2411 timepoints had AV-45 measurements. These participants were then divided into the four possible APOE profiles (See Figure 1).

**Figure 1:**
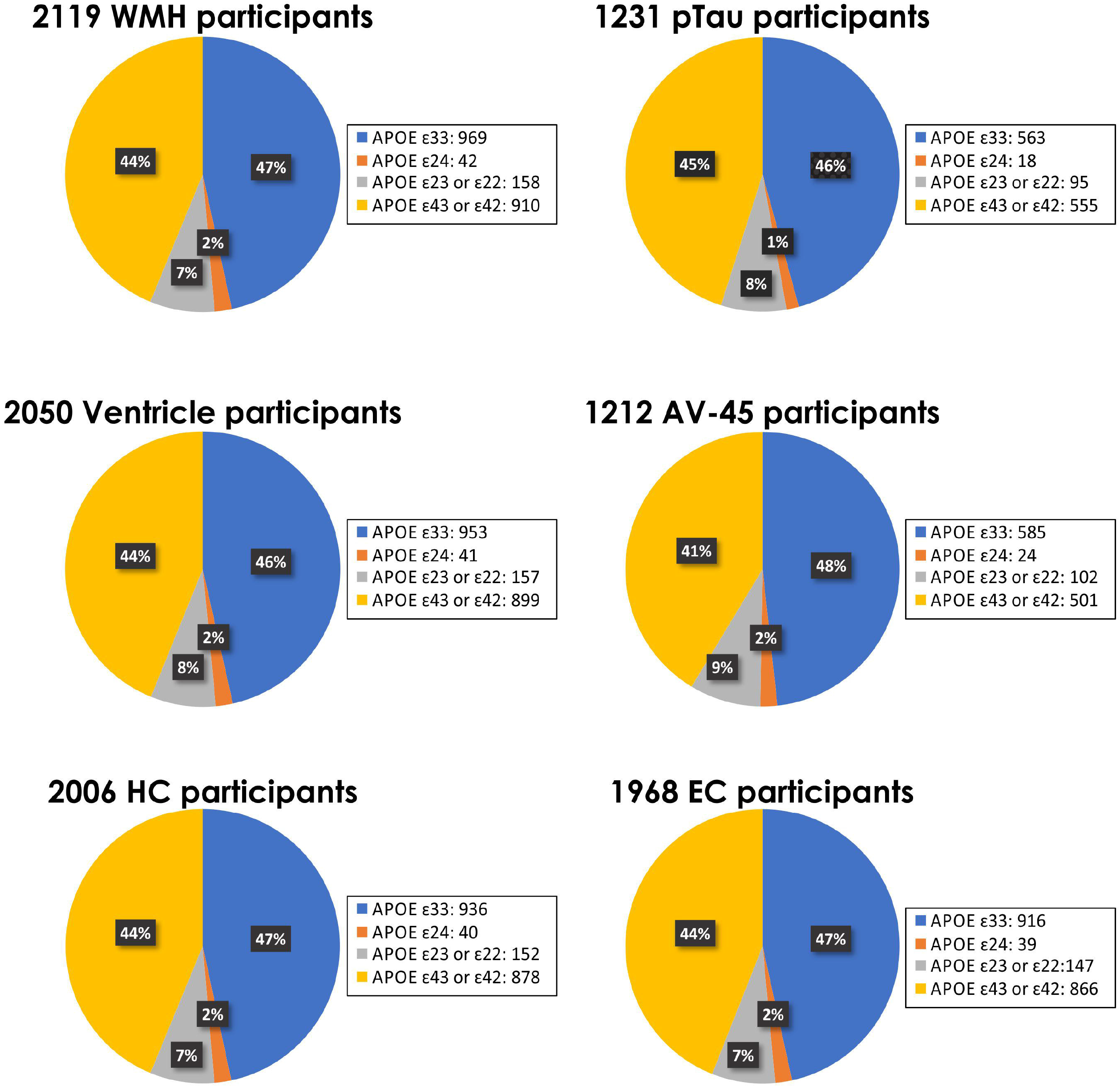
Proportion of participants in each analysis by APOE profile. *Notes:* Each plot presents the total number of participants with that pathology information. The percentage of the population with each APOsE genotype is also provided.

### 2.3 Structural MRI acquisition and processing

All longitudinal scans were downloaded from the ADNI website (see http://adni.loni.usc.edu/methods/mri-tool/mri-analysis/ for the detailed MRI acquisition protocol).T1w scans for each participant were pre-processed through our standard pipeline including noise reduction (28), intensity inhomogeneity correction (29), and intensity normalization into range [0-100]. The pre-processed images were then linearly (9 parameters: 3 translation, 3 rotation, and 3 scaling) (22) registered to the MNI-ICBM152-2009c average (30).

### 2.4 WMH measurements

A previously validated WMH segmentation technique was employed to generate participant WMH measurements (20). This technique has been validated in ADNI in which a library of manual segmentations based on 50 ADNI participants (independent of those studied here) was created. The technique has also been validated in other multi-center studies such as the Parkinson’s Markers Initiative (31) and the National Alzheimer’s Coordinating Center (32). WMHs were automatically segmented using the T1w contrasts, along with a set of location and intensity features obtained from a library of manually segmented scans in combination with a random forest classifier to detect the WMHs in new images (33,34). WMH load was defined as the volume of all voxels as WMH in the standard stereotaxic space (in mm^3^) and are thus normalized for head size. The volumes of the WMHs for frontal, parietal, temporal, and occipital lobes as well as the entire brain were calculated based on regional masks from the Hammers atlas (33,35). The quality of the registrations and WMH segmentations was visually verified by an experienced rater (author M.D.), blinded to diagnostic group.

### 2.5 pTau and AV-45 measurements

pTau and AV-45 measurements were obtained from ADNI. The pTau measurements were extracted from CSF samples obtained through lumbar punctures as described in the ADNI procedures manual. The pTau values were generated from the multiplex xMAP Luminex platform (Luminex Corp, Austin, TX, USA) with the INNO-BIA AlzBio3 kit (Innogenetics) (36,37). AV-45 PET imaging was performed within 2 weeks (before or after) the baseline clinical assessments for all participants with follow-up imaging at 2 years. Full description of procedures and processing has been previously described (38).

### 2.6 HC, EC, and ventricle measurements

Hippocampal, entorhinal cortex, and ventricle volumes were computed by ADNI using their standardized methods. Volumes were downloaded from the ADNI website.

### 2.7 Statistical Analysis

Analyses were performed using ‘R’ software version 4.0.5. WMH and ventricle volumes were log-transformed to achieve a more normal distribution. Linear mixed effects models were used to investigate the association between each pathology and the APOE groups. WMH load was examined for whole brain and frontal, temporal, parietal, and occipital regions. Regional WMH values (i.e., frontal, temporal, parietal, and occipital) were summed across the right and left hemispheres to obtain one score for each region. All continuous values (including log-transformed WMH volumes) were z-scored within the population prior to the analyses. All results were corrected for multiple comparisons using false discovery rate (FDR) of 0.05, p-values are reported as raw values with significance then determined by FDR correction (39).

The first set of linear regressions were completed to determine if baseline pathology measures differed between the different APOE profiles. Age, education, sex, and baseline diagnosis were included as covariates. Models were run separately for each dependent variable: WMH burden at each region, pTau, AV-45, ventricle volume, hippocampal volume, and entorhinal cortex volume.

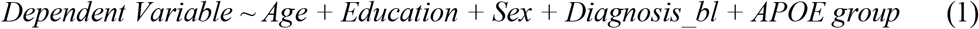

The second set of analyses included linear mixed effects models to determine if longitudinal change in pathology measures differed between the different APOE profiles. Age at baseline, education, sex, and baseline diagnosis were included as covariates. The interaction of interest was APOE group by TimeFromBaseline to examine if rate of change in pathology accumulation differed by APOE group. Longitudinal Models were run separately for each dependent variable including WMH burden at each region, pTau, AV-45, ventricle volume, hippocampal volume, and entorhinal cortex volume. In this model, participant ID was included as a categorical random effect.

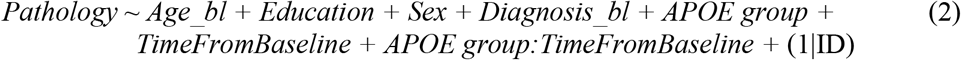

## 3. Results

As can be observed in Figure 2 and Table 1, group differences were observed in the baseline results. The APOE ε4 and ε2ε4 were associated with increased AV-45 compared to ε2 and ε3 (*t* belongs to [13.49 – 3.30], *p*<.001). The APOE ε4 was also associated with increased pTau compared to ε2 (*t*= 6.03, *p*<.001) and ε3 (*t=*9.82, *p*<.001). With respect to WMH burden, only the occipital region showed baseline differences, with ε4 having increased occipital WMH burden compared to ε2 (*t*= 3.11, *p*=.002) and ε3 (*t=* 2.35, *p* =.019). The ε2ε4 group had larger ventricles than ε3 (*t=* 2.46, *p* =.014) and ε4 (*t=* 2.49, *p* =.013). With respect to neurodegeneration, ε4 had smaller HC volumes than all other groups (*t* belongs to [2.46 – 6.06], *p*<.01), and smaller EC volumes compared to ε2 (*t*= 2.95, *p*=.003) and ε3 (*t*= 4.02, *p*<.001). No other APOE group differences were significant.

**Figure 2:**
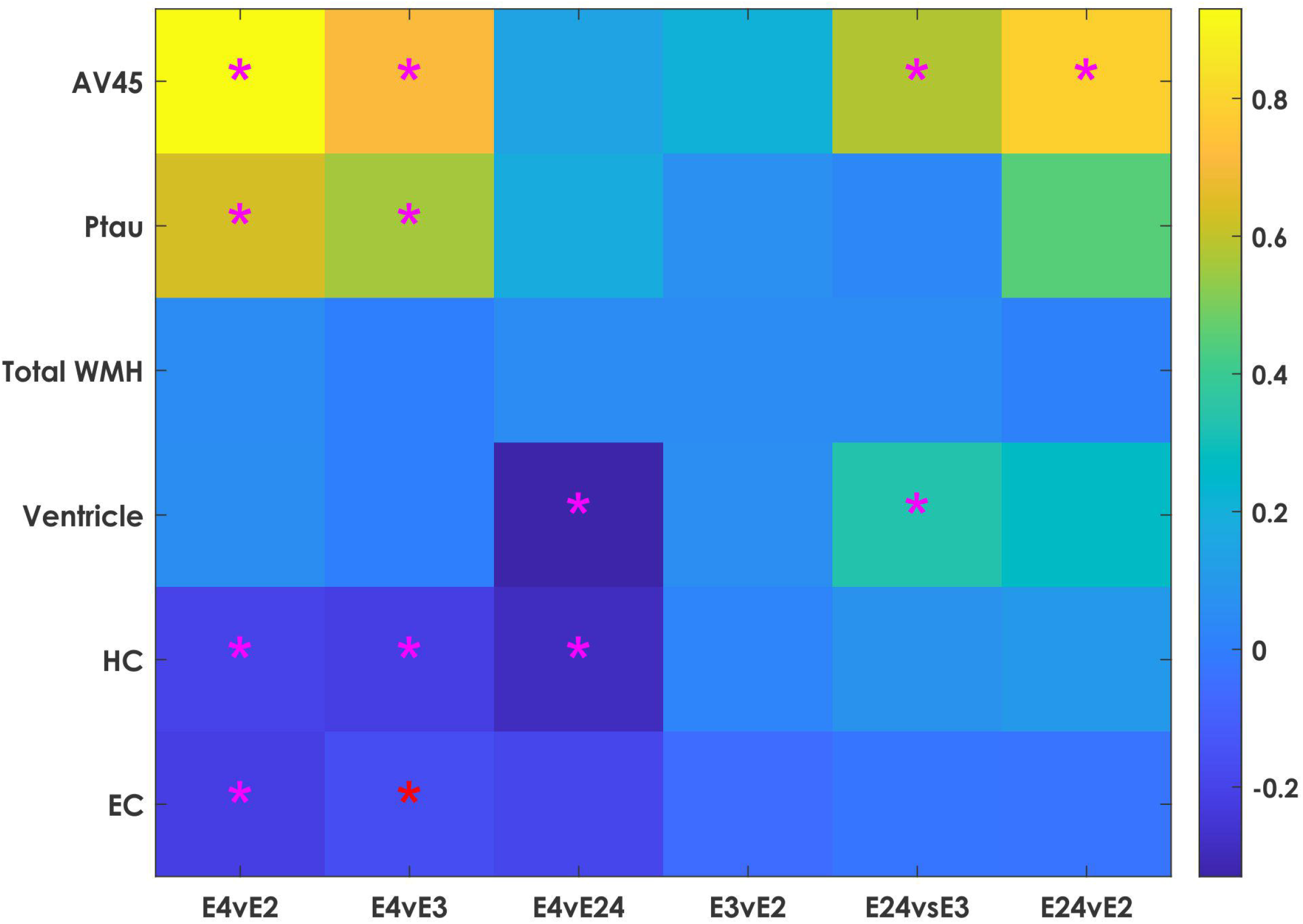
Colormap plots showing normalized beta estimates and an asterisk identifies statistically significant differences between the APOE profiles at baseline. *Notes*: Colormap showing baseline groups differences for each measure of pathology. Color values represent beta estimates of the z-scored values with significant differences marked by an asterisk. Each column represents a comparison between the two groups presented.

**Table 1:** Table showing estimates provided from the baseline linear regression.

| | $\epsilon 4 \vee \epsilon 2$ | $\epsilon 4 \vee \epsilon 3$ | $\epsilon 4 \vee \epsilon 24$ | $\epsilon 3 \vee \epsilon 2$ | $\epsilon 24 \vee \epsilon 3$ | $\epsilon 24 \vee \epsilon 2$ |
| --- | --- | --- | --- | --- | --- | --- |
| AV-45 | **$\beta=0.93$,**<br>**SE=0.09,**<br>**$t=9.90$,**<br>**$p<.001$** | **$\beta=0.72$,**<br>**SE=0.06,**<br>**$t=13.49$,**<br>**$p<.001$** | $\beta=0.14$,<br>SE=0.18,<br>$t=0.78$,<br>$p=.44$ | $\beta=0.21$,<br>SE=0.09,<br>$t=2.28$,<br>$p=.023$ | **$\beta=0.58$,**<br>**SE=0.18,**<br>**$t=3.30$,**<br>**$p=.001$** | **$\beta=0.79$,**<br>**SE=0.19,**<br>**$t=4.10$,**<br>**$p<.001$** |
| pTau | **$\beta=0.63$,**<br>**SE=0.10,**<br>**$t=6.03$,**<br>**$p<.001$** | **$\beta=0.56$,**<br>**SE=0.06,**<br>**$t=9.82$,**<br>**$p<.001$** | $\beta=0.18$,<br>SE=0.22,<br>$t=0.82$,<br>$p=.41$ | $\beta=0.06$,<br>SE=0.10,<br>$t=0.65$,<br>$p=.51$ | $\beta=0.38$,<br>SE=0.22,<br>$t=1.73$,<br>$p=.08$ | $\beta=0.45$,<br>SE=0.24,<br>$t=1.90$,<br>$p=.06$ |
| Occipital WMH | **$\beta=0.25$,**<br>**SE=0.08,**<br>**$t=3.10$,**<br>**$p=.002$** | **$\beta=0.10$,**<br>**SE=0.04,**<br>**$t=2.35$,**<br>**$p=.019$** | $\beta=0.05$,<br>SE=0.15,<br>$t=0.36$,<br>$p=.72$ | $\beta=0.15$,<br>SE=0.08,<br>$t=1.87$,<br>$p=.06$ | $\beta=0.05$,<br>SE=0.15,<br>$t=0.36$,<br>$p=.72$ | $\beta=0.20$,<br>SE=0.16,<br>$t=1.25$,<br>$p=.21$ |
| Ventricle | $\beta=0.06$,<br>SE=0.07,<br>$t=0.78$,<br>$p=.43$ | $\beta=0.01$,<br>SE=0.04,<br>$t=0.06$,<br>$p=.95$ | **$\beta=0.22$,**<br>**SE=0.13,**<br>**$t=2.46$,**<br>**$p=.014$** | $\beta=0.06$,<br>SE=0.07,<br>$t=0.84$,<br>$p=.40$ | **$\beta=0.33$,**<br>**SE=0.13,**<br>**$t=2.49$,**<br>**$p=.013$** | $\beta=-0.28$,<br>SE=0.15,<br>$t=-1.84$,<br>$p=.07$ |
| HC | **$\beta=-0.20$,**<br>**SE=0.07,**<br>**$t=-2.96$,**<br>**$p=.003$** | **$\beta=-0.22$,**<br>**SE=0.04,**<br>**$t=-6.06$,**<br>**$p<.001$** | **$\beta=-0.29$,**<br>**SE=0.11,**<br>**$t=-2.46$,**<br>**$p=.01$** | $\beta=-0.02$,<br>SE=0.06,<br>$t=-0.34$,<br>$p=.74$ | $\beta=-0.08$,<br>SE=0.12,<br>$t=-0.64$,<br>$p=.53$ | $\beta=-0.10$,<br>SE=0.13,<br>$t=-0.74$,<br>$p=.46$ |
| EC | **$\beta=-0.22$,**<br>**SE=0.07,**<br>**$t=-2.95$,**<br>**$p=.003$** | **$\beta=-0.16$,**<br>**SE=0.04,**<br>**$t=-4.02$,**<br>**$p<.001$** | $\beta=-0.19$,<br>SE=0.13,<br>$t=-1.39$,<br>$p=.16$ | $\beta=-0.06$,<br>SE=0.07,<br>$t=-0.80$,<br>$p=.43$ | $\beta=-0.25$,<br>SE=0.13,<br>$t=-0.18$,<br>$p=.85$ | $\beta=-0.33$,<br>SE=0.15,<br>$t=-0.23$,<br>$p=.82$ |

Notes: Occipital WMH burden is provided as opposed to total because it was the only WMH measure significant between groups at baseline. WMH = white matter hyperintensity. HC = hippocampus. EC = entorhinal cortex. Bold values are those that remained significant after correction for multiple comparisons.

Longitudinal results can be observed in Figure 3 and Table 2. The APOE ε4 group had increased rate of AV-45 change compared to ε2 (*t*= 4.91, *p*<.001) and ε3 (*t*= 3.83, *p*<.001), and ε3 had increased rate of AV-45 change compared to ε2 (*t*= 2.75, *p*=.006). Interestingly, the ε4 group had a smaller rate of pTau accumulation than the ε3 group (*t*= -4.76, *p*<.001). Rate of total WMH accumulation differed between all groups (*t* belongs to [10.42 – 2.61], *p*<.001), except ε2ε4 vs. ε4 and ε2ε4 vs. ε3. Similar results were obtained for regional WMH burden rates (see Table 3). Rate of change for ventricle (*t* belongs to [20.92 – 2.57], *p*<.01) and hippocampal volume (*t* belongs to [17.59 – 2.48], *p*<.01) significantly differed between all groups. The ε4 group had increased EC atrophy over time compared to ε2 (*t*= 7.42, *p*<.001) and ε3 (*t*= 9.87, *p*<.001), and the ε2ε4 had increased EC atrophy over time compared to ε2 (*t*= 2.43, *p*=.015).

**Figure 3:**
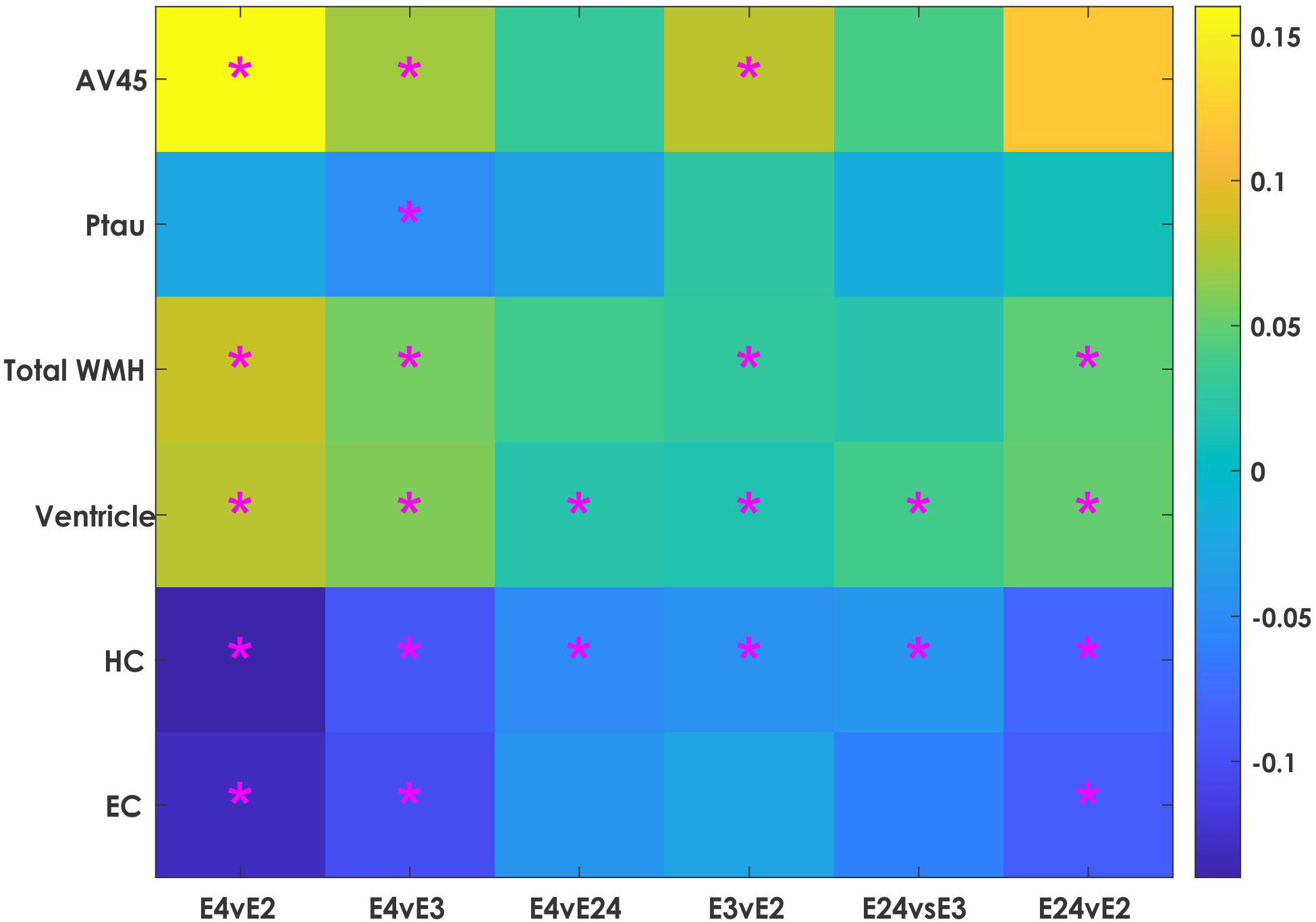
Colormap plots showing normalized beta estimates and an asterisk identifies statistically significant differences between the APOE profiles over time. *Notes*: Colormap showing the interaction of APOE profile and time from baseline to reflect rate of change group differences for each measure of pathology. Color values represent the beta estimates of the z-scored values with significant differences marked by an asterisk. Each column represents a comparison between the two groups presented.

**Table 2:** Table showing estimates provided from the longitudinal linear mixed effects models interaction between APOE profile and time.

| | $\epsilon 4v\epsilon 2$ | $\epsilon 4v\epsilon 3$ | $\epsilon 4v\epsilon 24$ | $\epsilon 3v\epsilon 2$ | $\epsilon 24v\epsilon 3$ | $\epsilon 24v\epsilon 2$ |
| --- | --- | --- | --- | --- | --- | --- |
| AV-45 | **$\beta=0.16$,**<br>**SE=0.03,**<br>**$t=4.91$,**<br>**$p<.001$** | **$\beta=0.07$,**<br>**SE=0.02,**<br>**$t=3.83$,**<br>**$p<.001$** | $\beta=0.03$,<br>SE=0.06,<br>$t=0.52$,<br>$p=.60$ | **$\beta=0.08$,**<br>**SE=0.03,**<br>**$t=2.75$,**<br>**$p=.006$** | $\beta=0.04$,<br>SE=0.06,<br>$t=0.66$,<br>$p=.51$ | $\beta=0.12$,<br>SE=0.07,<br>$t=1.85$,<br>$p=.064$ |
| pTau | $\beta=0.02$,<br>SE=0.02,<br>$t=1.23$,<br>$p=.22$ | **$\beta=-0.05$,**<br>**SE=0.01,**<br>**$t=-4.76$,**<br>**$p<.001$** | $\beta=0.03$,<br>SE=0.02,<br>$t=1.27$,<br>$p=.21$ | $\beta=0.03$,<br>SE=0.02,<br>$t=1.62$,<br>$p=.11$ | $\beta=0.38$,<br>SE=0.22,<br>$t=1.73$,<br>$p=.08$ | $\beta=0.45$,<br>SE=0.24,<br>$t=1.90$,<br>$p=.06$ |
| Total WMH | **$\beta=0.08$,**<br>**SE=0.01,**<br>**$t=9.29$,**<br>**$p<.001$** | **$\beta=0.06$,**<br>**SE=0.01,**<br>**$t=10.42$,**<br>**$p<.001$** | $\beta=0.03$,<br>SE=0.01,<br>$t=2.13$,<br>$p=.03$ | **$\beta=0.03$,**<br>**SE=0.01,**<br>**$t=3.24$,**<br>**$p=.001$** | $\beta=0.02$,<br>SE=0.02,<br>$t=1.17$,<br>$p=.24$ | **$\beta=0.05$,**<br>**SE=0.02,**<br>**$t=2.61$,**<br>**$p=.009$** |
| Ventricle | **$\beta=0.08$,**<br>**SE=0.01,**<br>**$t=16.04$,**<br>**$p<.001$** | **$\beta=0.06$,**<br>**SE=0.01,**<br>**$t=20.92$,**<br>**$p<.001$** | **$\beta=0.02$,**<br>**SE=0.01,**<br>**$t=2.57$,**<br>**$p=.010$** | **$\beta=0.02$,**<br>**SE=0.01,**<br>**$t=3.53$,**<br>**$p<.001$** | **$\beta=0.04$,**<br>**SE=0.01,**<br>**$t=3.94$,**<br>**$p<.001$** | **$\beta=0.05$,**<br>**SE=0.01,**<br>**$t=5.25$,**<br>**$p<.001$** |
| HC | **$\beta=-0.14$,**<br>**SE=0.01,**<br>**$t=-15.37$,**<br>**$p<.001$** | **$\beta=-0.09$,**<br>**SE=0.01,**<br>**$t=-17.59$,**<br>**$p<.001$** | **$\beta=-0.05$,**<br>**SE=0.02,**<br>**$t=-3.19$,**<br>**$p=.001$** | **$\beta=-0.04$,**<br>**SE=0.01,**<br>**$t=-5.13$,**<br>**$p<.001$** | **$\beta=-0.04$,**<br>**SE=0.02,**<br>**$t=-2.48$,**<br>**$p=.013$** | **$\beta=-0.08$,**<br>**SE=0.02,**<br>**$t=-4.73$,**<br>**$p<.001$** |
| EC | **$\beta=-0.14$,**<br>**SE=0.02,**<br>**$t=-7.42$,**<br>**$p<.001$** | **$\beta=-0.11$,**<br>**SE=0.01,**<br>**$t=-9.87$,**<br>**$p<.001$** | $\beta=-0.05$,<br>SE=0.03,<br>$t=-1.43$,<br>$p=.15$ | $\beta=0.03$,<br>SE=0.02,<br>$t=1.53$,<br>$p=.13$ | $\beta=-0.06$,<br>SE=0.03,<br>$t=-1.84$,<br>$p=.065$ | **$\beta=-0.09$,**<br>**SE=0.04,**<br>**$t=-2.43$,**<br>**$p=.015$** |

Notes: WMH = white matter hyperintensity. HC = hippocampus. EC = entorhinal cortex. Bold values are those that remained significant after correction for multiple comparisons.

**Table 3:** Table showing estimates provided from the longitudinal linear mixed effects modelling the interaction between APOE profile and time for regional WMH burden.

| | $\epsilon 4 \vee \epsilon 2$ | $\epsilon 4 \vee \epsilon 3$ | $\epsilon 4 \vee \epsilon 24$ | $\epsilon 3 \vee \epsilon 2$ | $\epsilon 24 \vee \epsilon 3$ | $\epsilon 24 \vee \epsilon 2$ |
| --- | --- | --- | --- | --- | --- | --- |
| Frontal | $\beta=0.07$,<br>$SE=0.01$,<br>$t=8.09$,<br>$p<.001$ | $\beta=0.05$,<br>$SE=0.01$,<br>$t=9.11$,<br>$p<.001$ | $\beta=0.01$,<br>$SE=0.02$,<br>$t=0.65$,<br>$p=.52$ | $\beta=0.02$,<br>$SE=0.01$,<br>$t=2.80$,<br>$p=.005$ | $\beta=0.04$,<br>$SE=0.02$,<br>$t=2.25$,<br>$p=.024$ | $\beta=0.06$,<br>$SE=0.02$,<br>$t=3.39$,<br>$p<.001$ |
| Temporal | $\beta=0.07$,<br>$SE=0.01$,<br>$t=6.41$,<br>$p<.001$ | $\beta=0.03$,<br>$SE=0.01$,<br>$t=4.76$,<br>$p<.001$ | $\beta=0.05$,<br>$SE=0.02$,<br>$t=2.68$,<br>$p<.001$ | $\beta=0.04$,<br>$SE=0.01$,<br>$t=3.74$,<br>$p<.001$ | $\beta=0.02$,<br>$SE=0.02$,<br>$t=1.20$,<br>$p=.23$ | $\beta=0.01$,<br>$SE=0.02$,<br>$t=0.67$,<br>$p=.50$ |
| Parietal | $\beta=0.08$,<br>$SE=0.01$,<br>$t=9.09$,<br>$p<.001$ | $\beta=0.05$,<br>$SE=0.01$,<br>$t=10.23$,<br>$p<.001$ | $\beta=0.05$,<br>$SE=0.02$,<br>$t=2.91$,<br>$p=.003$ | $\beta=0.03$,<br>$SE=0.01$,<br>$t=3.14$,<br>$p=.001$ | $\beta=0.01$,<br>$SE=0.02$,<br>$t=0.33$,<br>$p=.75$ | $\beta=0.03$,<br>$SE=0.02$,<br>$t=1.78$,<br>$p=.07$ |
| Occipital | $\beta=0.13$,<br>$SE=0.01$,<br>$t=9.67$,<br>$p<.001$ | $\beta=0.08$,<br>$SE=0.01$,<br>$t=10.35$,<br>$p<.001$ | $\beta=0.07$,<br>$SE=0.02$,<br>$t=2.62$,<br>$p=.009$ | $\beta=0.05$,<br>$SE=0.01$,<br>$t=3.68$,<br>$p<.001$ | $\beta=0.02$,<br>$SE=0.02$,<br>$t=0.65$,<br>$p=.51$ | $\beta=0.06$,<br>$SE=0.03$,<br>$t=2.34$,<br>$p=.019$ |

Notes: Bold values are those that remained significant after correction for multiple comparisons.

## 4. Discussion

Over the last several decades, an abundance of research has attempted to identify early risk factors of AD in order to mitigate cognitive decline and even prevent disease progression. One of the important risk factors of cognitive aging (40) and conversion to AD is presence of the APOE ε4 allele (41). The relationship between the ε2 allele and ε2ε4 genotype with cognitive functioning and AD progression remains more elusive than with ε4. The current study helps improve our understanding of how different APOE profiles are associated with neurodegeneration and brain pathology. At baseline, ε4 exhibited increased pTau compared to ε2 and ε3 and smaller HC and EC volumes than all other APOE profiles. The ε2ε4 group exhibited smaller ventricles than ε3 and ε4. The ε2 group exhibited less amyloid than all other APOE profiles, while ε4 and ε2ε4 had more amyloid compared to ε3. Longitudinally, many more APOE profile differences were apparent. Ventricular enlargement and HC atrophy significantly differed between all profiles. Furthermore, ε2 was observed to have slower WMH accumulation compared to all other profiles, while E4 exhibited faster accumulation compared to ε3. The ε4 group exhibited increased EC atrophy compared to ε3 and ε2, and ε2ε4 exhibited more atrophy than ε2. Amyloid increased faster in the ε3 group compared to ε3 and ε2, while ε2 amyloid progressed slower than ε3.

Consistent with previous findings indicating an association between ε4 and increased Aβ(6), we observed that ε4 group exhibited elevated Aβ levels compared to ε2 and ε3 at baseline and had greater change over time. At baseline, the ε2ε4 group also exhibited increased Aβ levels compared to ε2 and ε3, but they did not exhibit an increased rate of change longitudinally. The protective effect of the ε2 also resulted in reduced accumulation of Aβ also compared to those with ε3. These findings are consistent with past reports that the ε4 is associated with increased Aβ while the ε2 is associated with lower Aβ (11,12). Additionally, this study observed similar ε4 and ε2 effects in rate of accumulation of Aβ, indicating the detrimental effect of ε4 and protective effect of ε2 on rate of amyloid accumulation. The finding of increased baseline ε2ε4 Aβ, but lack of longitudinal differences indicates that the change over time in this group is similar to that of all other groups. The heightened Aβ at baseline compared to ε2 and ε3 may reduce resiliency to other brain changes observed longitudinally.

Interestingly, although ε4 had increased pTau levels at baseline compared to ε2 and ε3, this group did not have increased rates of pTau levels longitudinally. No other pTau differences between the groups were observed at baseline or longitudinally. The minimal pTau differences between APOE profiles may be because of the inclusion of all diagnostic groups in our analyses. Previous research has observed that the relationship between APOE status and tau is found only in the presence of Aβ (10), therefore including some amyloid negative normal controls may reduce the longitudinal associations. This finding may be a limitation of the current study as the sample size is not large enough to examine APOE profiles within each diagnostic group individually (i.e., cognitively healthy older adults, MCI, and AD).

WMH burden was measured as total brain WMH volume as well as regionally (at frontal, temporal, parietal, and occipital regions). At baseline, only occipital WMH burden showed differences based on APOE profile. More specifically, ε4 had increased occipital WMHs over both ε2 and ε3. The ε4 profile showed increased rates of WMH accumulation compared to ε2 and ε3 for total burden and for all regions, and more than ε2ε4 at the temporal, parietal, and occipital regions. Previous research has observed that parietal and occipital WMHs are the most prominent areas associated with WMH volume observed in AD (42,43). Therefore, people with the ε4 profile are exhibiting WMH burden that is associated with progression to dementia. The ε2 profile showed lower rates of WMH accumulation compared to ε3 for total burden and at all regions, and compared to ε2ε4 for total WMH accumulation and frontal and occipital regions. These findings are consistent with cross-sectional findings showing the protective effects of ε2 (23) and detrimental effects of ε4 (24,25). Extending on this research, the current study also observed that the ε2 offers protection against WMH accumulation over time. With respect to the ε2ε4, this group showed less WMH accumulation compared to ε4, no difference compared to ε3, but more than ε2 at total, frontal, and occipital regions. They showed a similar pattern of WMH change to that of ε3, indicating that the ε2ε4 profile provides a neutral effect on WMH burden.

Examination of overall neurodegeneration and overall atrophy was completed using ventricle volume. At baseline, the only ventricle differences observed were limited to ε2ε4 having larger ventricles compared to both ε3 and ε4. This slightly larger ventricle volume at baseline may be associated with the limited sample size of the ε2ε4. This baseline difference was followed by the ε2ε4 having increased ventricle volume over time compared to ε2 and ε3, but slightly lower rate of change compared to ε4. These findings suggest that the ε2ε4 genotype has a detrimental risk towards overall atrophy as measured by ventricle volumes. The ε4 profile had increased ventricle volume rate of change compared to both ε3 and ε2, and ε2 had less ventricle volume increases than ε3. That is, ε4 has a negative influence on ventricle volume while ε2 offers protection towards minimizing ventricle size. Despite inconsistent results in the literature on the relationship between ventricle size and APOE status (14), our findings suggest a strong relationship between APOE and ventricle volume.

Both baseline and longitudinal rate of hippocampal and entorhinal cortex volume were observed to be associated with APOE profile. Consistent with previous findings, at baseline the ε4 profile had increased hippocampal atrophy compared to all other groups (16), and increased entorhinal cortex atrophy compared to ε2 and ε3 (14). Rate of change of hippocampal volume was significantly different between all APOE profiles. The ε2ε4 exhibited increased hippocampal atrophy over time compared to ε2 and ε3, but lower rate of change compared to ε4. These findings suggest that the ε2ε4 genotype has a detrimental risk towards hippocampal atrophy, a known marker of AD disease staging (44). The ε4 profile had increased hippocampal atrophy over time compared to both ε3 and ε2, and ε2 had less hippocampal atrophy increases than ε3. That is, ε4 has a negative influence, while ε2 offers protection towards minimizing hippocampal atrophy that occurs over time. Rate of atrophy change in the entorhinal cortex was increased in ε4 compared to ε2 and ε3, and ε2ε4 compared to ε2. As the ε2ε4 did not differ from ε3 or ε4 but was slightly increased compared to, we can interpret this finding as the ε2ε4 profile exhibiting an intermediate rate of change between ε3 and ε4.

It should be noted that a weakness of the current study is the use of only ADNI data. This sample is highly educated and lacks diversity. Our participants had an average education of 16 years and were mainly white individuals (93% of the sample), which may reduce generalizability to more representative samples. Given that previous research has observed that the relationship between AD and APOE differs based on race (45), it is imperative that future research explore the longitudinal relationship between APOE status and pathology in other races. The longitudinal nature of this project is a major strength, as it improves our ability to draw causal relationships between APOE status and neuropathology. This study provides an in-depth analysis of both the protective and detrimental effects APOE can have on AD related pathology.

The strong associations observed between APOE and pathology in this study show the importance of how genetic factors influence structural brain changes. These findings offer clarification on the protective effects that ε2 offers and the detrimental effects from the ε4 towards neurodegeneration and pathologies. Furthermore, the observation of the detrimental effect of ε2ε4 on both ventricle volume and hippocampal atrophy change over time is a novel result that may improve treatments and interventions. From a clinical standpoint, previous work has shown that APOE-specific targeted interventions (46) can help mitigate cognitive decline in people with ε4 status, and may offer greater chances of successful techniques to prevent AD. Given that people who have the ε2ε4 genotype can expect to have increased atrophy, they must be included (alongside those with an ε4 profile) in targeted interventions to reduce brain changes that occur with AD.

## Data Availability

All data produced are available online at adni.loni.usc.edu

## Declarations

## Acknowledgments

Data collection and sharing for this project was funded by the Alzheimer’s Disease Neuroimaging Initiative (ADNI) (National Institutes of Health Grant U01 AG024904) and DOD ADNI (Department of Defense award number W81XWH-12-2-0012). ADNI is funded by the National Institute on Aging, the National Institute of Biomedical Imaging and Bioengineering, and through generous contributions from the following: AbbVie, Alzheimer’s Association; Alzheimer’s Drug Discovery Foundation; Araclon Biotech; BioClinica, Inc.; Biogen; Bristol-Myers Squibb Company; CereSpir, Inc.; Cogstate; Eisai Inc.; Elan Pharmaceuticals, Inc.; Eli Lilly and Company; EuroImmun; F. Hoffmann-La Roche Ltd and its affiliated company Genentech, Inc.; Fujirebio; GE Healthcare; IXICO Ltd.; Janssen Alzheimer Immunotherapy Research & Development, LLC.; Johnson & Johnson Pharmaceutical Research & Development LLC.; Lumosity; Lundbeck; Merck & Co., Inc.; Meso Scale Diagnostics, LLC.; NeuroRx Research; Neurotrack Technologies; Novartis Pharmaceuticals Corporation; Pfizer Inc.; Piramal Imaging; Servier; Takeda Pharmaceutical Company; and Transition Therapeutics. The Canadian Institutes of Health Research is providing funds to support ADNI clinical sites in Canada. Private sector contributions are facilitated by the Foundation for the National Institutes of Health (www.fnih.org). The grantee organization is the Northern California Institute for Research and Education, and the study is coordinated by the Alzheimer’s Therapeutic Research Institute at the University of Southern California. ADNI data are disseminated by the Laboratory for Neuro Imaging at the University of Southern California.

## Ethics Approval

The study received ethical approval from the review boards of all participating institutions.

## Consent Statement

Written informed consent was obtained from participants or their study partner.

## Authors Contributions

C.M, M.D, F.K, and D.L.C. were involved with the conceptualization and design of the work. C.M and M.D. completed analysis and C.M, M.D, F.K, and D.L.C. were involved with data interpretation. C.M wrote the manuscript. and C.M, M.D, F.K, and D.L.C. revised and approved the submitted version.

## Data availability and materials

The datasets generated and/or analysed during the current study are available in the ADNI repository, adni.loni.usc.edu.

## List of abbreviations

(AD): Alzheimer’s disease
(ADNI): Alzheimer’s Disease Neuroimaging Initiative
(APOE): apolipoprotein
(Aβ): beta-amyloid
(CSVD): cerebral small vessel disease
(CDR): clinical dementia rating
(EC): entorhinal cortex
(FDR): false discovery rate
(HC): hippocampus
(MMSE): Mini Mental Status Examination
(WMHs): white matter hyperintensities

